# Genome sequencing and analysis of an emergent SARS-CoV-2 variant characterized by multiple spike protein mutations detected from the Central Visayas Region of the Philippines

**DOI:** 10.1101/2021.03.03.21252812

**Authors:** Francis A. Tablizo, Kenneth M. Kim, Carlo M. Lapid, Marc Jerrone R. Castro, Maria Sofia L. Yangzon, Benedict A. Maralit, Marc Edsel C. Ayes, Eva Maria Cutiongco-de la Paz, Alethea R. De Guzman, Jan Michael C. Yap, Jo-Hannah S. Llames, Shiela Mae M. Araiza, Kris P. Punayan, Irish Coleen A. Asin, Candice Francheska B. Tambaoan, Asia Louisa U. Chong, Karol Sophia Agape R. Padilla, Rianna Patricia S. Cruz, El King D. Morado, Joshua Gregor A. Dizon, Razel Nikka M. Hao, Arianne A. Zamora, Devon Ray Pacial, Juan Antonio R. Magalang, Marissa Alejandria, Celia Carlos, Anna Ong-Lim, Edsel Maurice Salvaña, John Q. Wong, Jaime C. Montoya, Maria Rosario Singh-Vergeire, Cynthia P. Saloma

## Abstract

The emergence of SARS-CoV-2 variants of concern such as the B.1.1.7, B.1.351 and the P.1 have prompted calls for governments worldwide to increase their genomic biosurveillance efforts. Globally, quarantine and outbreak management measures have been implemented to stem the introduction of these variants and to monitor any emerging variants of potential clinical significance domestically. Here, we describe the emergence of a new SARS-CoV-2 lineage, mainly from the Central Visayas region of the Philippines. This emergent variant is characterized by 13 lineage-defining mutations, including the co-occurrence of the E484K, N501Y, and P681H mutations at the spike protein region, as well as three additional radical amino acid replacements towards the C-terminal end of the said protein. A three-amino acid deletion at positions 141 to 143 (LGV141_143del) in the spike protein was likewise seen in a region preceding the 144Y deletion found in the B.1.1.7 variant. A single amino acid replacement, K2Q, at the N-terminus of ORF8 was also shared by all 33 samples sequenced. The mutation profile of this new virus variant warrants closer investigation due to its potential public health implications. The current distribution of this emergent variant in the Philippines and its transmission are being monitored and addressed by relevant public health agencies to stem its spread in nearby islands and regions in the country.

## INTRODUCTION

On March 11, 2020, the coronavirus disease 2019 (COVID-19), caused by the Severe Acute Respiratory Syndrome Coronavirus 2 (SARS-CoV-2), was declared by the World Health Organization (WHO) as a global pandemic, causing over 115 million infections and over 2.5 million deaths as of March 2, 2021. The global uncontrolled spread of SARS-CoV-2 has given rise to various viral lineages circulating worldwide, with at least three (3) variants being flagged as variants of concern.

Early in December 2020, the United Kingdom reported the emergence of the B.1.1.7 variant *(20I/501Y.V1 Variant of Concern (VOC) 202012/0)*, defined by 17 amino acid changes, including eight (8) mutations in the spike protein (Rambaut et al., 2020). In addition to the B.1.1.7, two (2) more variants of concern were reported: the B.1.351 lineage (South African Variant *20H/501Y.V2*) and the more recent P.1 lineage (*20J/501Y.V3*) of Brazilian origin. All three lineages of concern share the spike N501Y mutation, whereas the P.1 and B.1.351 lineages additionally share two (2) other mutations in the spike protein (K417N/T and E484K) (COVID-19 Genomics UK Consortium [CoG-UK], 2021). Even though the set of mutations/deletions found in the B.1.1.7, B.1.351, and P.1 lineages appear to have emerged independently, these viruses have all been associated with a rapid increase in cases in their respective places of origin (Faria et al., 2021; Rambaut et al., 2020a; Tegally et al., 2020). The WHO has encouraged countries globally to increase routine systematic sequencing of SARS-CoV-2 viruses to better understand SARS-CoV-2 transmission and to monitor possible emergence of new variants (World Health Organization [WHO], 2021).

The Philippines is among the countries adversely hit by the COVID-19 pandemic, with over 580,000 confirmed cases and over 12,000 deaths attributed to the disease as of March 2, 2021. Although border restrictions are in place to prevent the spread of the virus, local and international travel is still ongoing, primarily for returning overseas Filipinos (ROFs) from various countries around the world. The announcement of the UK variant last December 2020 prompted the government to conduct a nationwide genomic surveillance program to detect SARS-CoV-2 variants, particularly among ROFs in their ports of entry, as well as from local case clusters in different regions of the archipelago.

In this study, we report the detection of 33 Philippine SARS-CoV-2 cases that were found to share 13 lineage-defining mutations that may have potential biological significance, including the co-occurrence of the E484K, N501Y, and P681H amino acid replacements at the spike protein region. The mutation profile observed for this set of viruses has not been described in currently known SARS-CoV-2 lineages of concern and were detected primarily from the Central Visayas Region of the country during a period that coincides with a recorded surge in COVID-19 cases in the area.

## RESULTS

The Philippine Department of Health, through its surveillance efforts, monitors and analyzes COVID-19 case data since the start of the pandemic. A decline in cases were seen after cases peaked in August of 2020. From January 21, the Department saw an increase in cases particularly in key areas in the country, including Central Visayas. While an increase in cases was not unexpected given the increased mobility, crowding, and gatherings during the holiday season, the cases in Central Visayas continued to rise as other areas started to plateau around February (**Figure 1**). To further enrich the surveillance data gathered from this area, samples from Central Visayas were prioritized for whole genome sequencing of the virus. In total, we sequenced 60 samples from this region.

**Figure 1.**
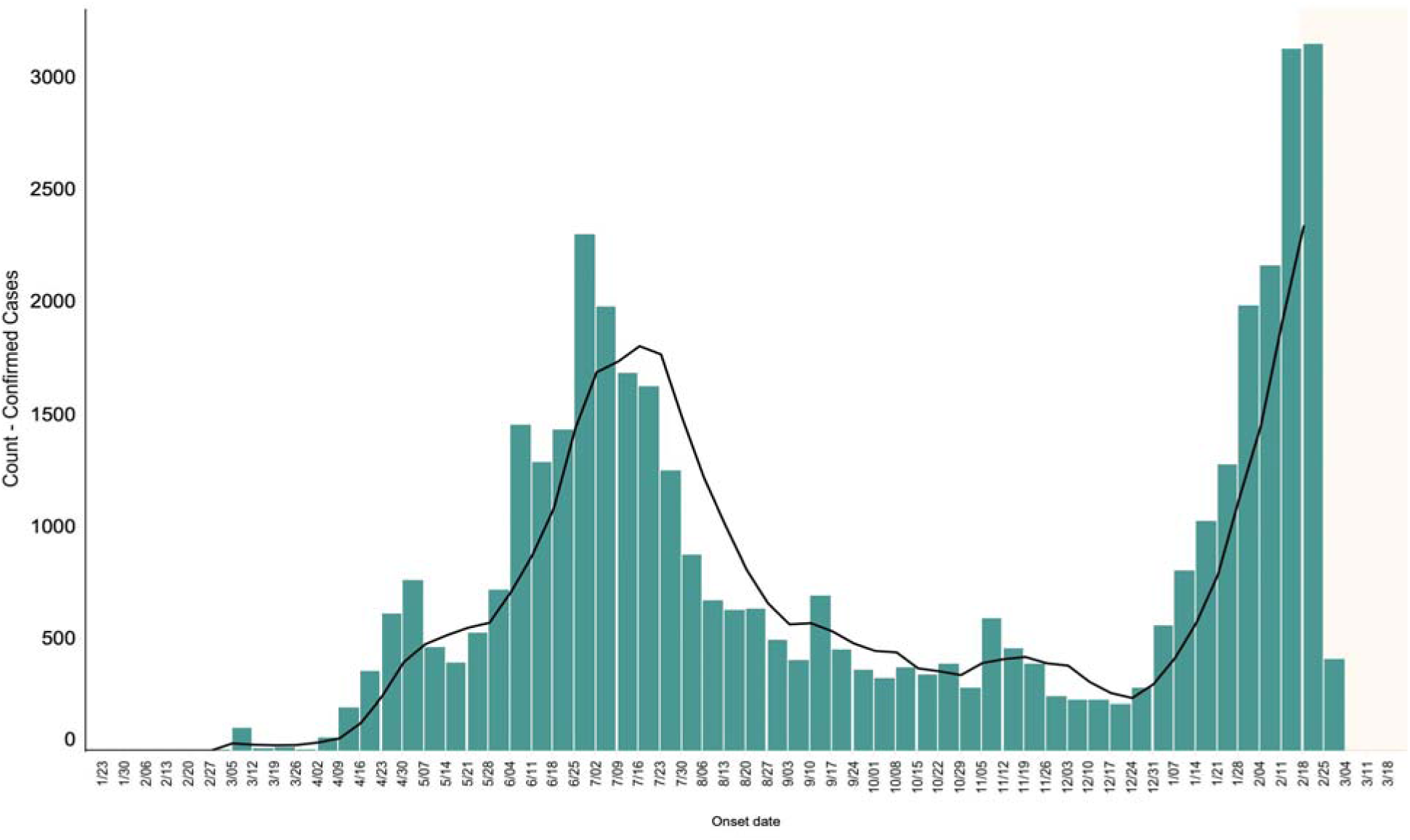
Epidemic curve in Central Visayas, Philippines. The figure above shows the distribution of reported COVID-19 confirmed cases from the Central Visayas region as to their date of onset of illness as of 02 March 2021. A steep rise in cases have been reported since the January 21, 2021.

We report here the detection of 33 Philippine SARS-CoV-2 infections caused by viruses initially classified to be under lineage B.1.1.28, but were found to have a co-occurrence of the spike protein E484K, N501Y, and P681H mutations. The samples were collected from January 16 to February 02, 2021 and most were among the sequenced samples from Central Visayas, with a few others from laboratory submissions from the National Capital Region (NCR) and Davao Region. **Table 1** shows the breakdown of sample counts per region of laboratory facility.

**Table 1.** Regions in the Philippines where the emergent SARS-CoV-2 variant was detected.

| Region of Laboratory Facility | # Cases | Date of Collection | Remarks |
| --- | --- | --- | --- |
| Central Visayas | 29 | January 31 – February 02, 2021 | Mostly from the Island of Cebu |
| National Capital Region | 3 | January 16 – February 01, 2021 | Earliest recorded collection |
| Davao Region | 1 | January 23, 2021 |  |

The three (3) co-occurring mutations have all been identified as significant SNPs in lineages of concern. The E484K and N501Y co-occur in the P.1 lineage (Brazilian Variant) and the B.1.351 lineage (South African Variant). In addition, N501Y and P681H are both defining single nucleotide polymorphisms (SNPs) for the B.1.1.7 lineage (UK Variant) (CoG-UK, 2021). However, a profile of other high-frequency mutations in the Philippine samples indicate that they do not share any of the other defining SNPs from any of these lineages (**Figure 2**) (Faria et al., 2021; Rambaut et al., 2020a; Tegally et al., 2020). All the samples also carry the globally prevalent spike D614G mutation.

**Figure 2.**
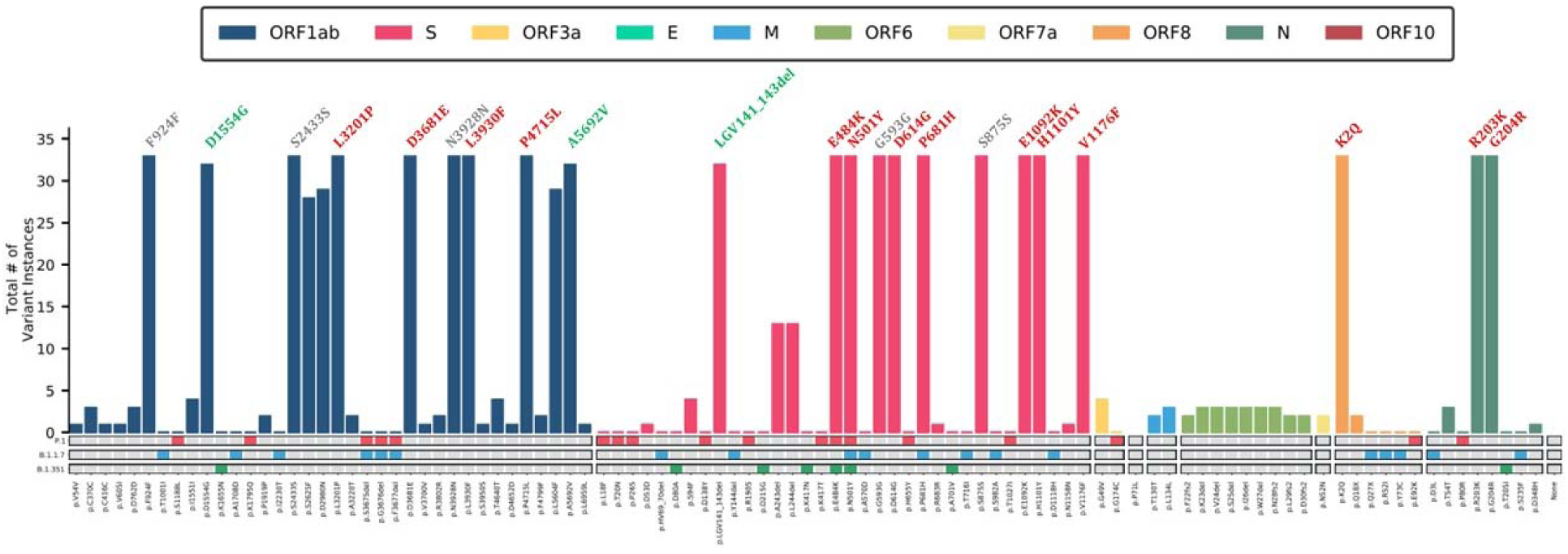
Mutation profile of 33 Philippine SARS-CoV-2 samples comprising an emergent viral lineage. A total of 14 amino acid replacements were observed in all samples (labeled in red), including seven (7) spike protein mutations. Among the spike protein mutations, four (4) have been previously associated with lineages of concern (*i.e*., E484K, N501Y, D614G, and P681H) while three (3) additional replacements were observed towards the C-terminal region of the protein (*i.e*., E1092K, H1101Y, and V1176F). Interestingly, a single amino acid replacement at the N-terminus of ORF8 (*i.e*., K2Q) was also found in all samples. Three (3) other mutations were seen in 32 of the 33 samples (labeled in green), including a three-amino acid deletion at the spike protein positions 141 to 143. Lastly, five (5) synonymous mutations (labeled in gray) were also detected in all of the cases. For comparison, the defining mutations for the P.1, B.1.1.7, and B.1.351 lineages of concern are marked below in red, blue, and green boxes, respectively. None of the samples exhibit mutation profiles similar to any of the three (3) known lineages of concern.

Apart from the spike protein mutations previously associated with SARS-CoV-2 lineages of concern, ten additional amino acid replacements were also observed in the 33 Philippine cases, including the following: L3201P, D3681E, L3930F and P4715L in the ORF1ab polyprotein; E1092K, H1101Y and V1176F in the spike protein; K2Q in the ORF8 protein product; and R203K and G204R in the nucleocapsid protein. Three (3) other mutations were found in 32 of the 33 Philippine samples, which include the D1554G and A5692V amino acid replacements at the ORF1ab polyprotein, as well as a three-amino acid deletion at the spike protein at positions 141 to 143 (LGV141_143del). Five (5) synonymous mutations were also seen: F924F, S2433S and N3928N in ORF1ab; and spike protein G593G and S875S. A list of the commonly observed mutations for this viral group is shown in **Table 2** and the designated signature mutations are also indicated. The significance of these mutations, particularly the combination of mutations in the spike and other regions of the viral genome, on the transmissibility, pathogenicity, and immunogenicity of this emergent variant remains to be studied.

**Table 2.** Mutations observed in at least 32 of the 33 Philippine SARS-CoV-2 samples comprising an emergent virus variant. The column “Signature” signifies the mutations found in this group of viruses that are likely to have functional implications or are not prevalent in other lineages.

| Affected Gene | Nucleotide Change | Amino Acid Change | # Cases | Signature |
| --- | --- | --- | --- | --- |
| **Non-synonymous Mutations** |  |  |  |  |
| ORF1ab | 4962A>G | D1554G | 32 | Yes |
| ORF1ab | 9867T>C | L3201P | 33 | Yes |
| ORF1ab | 11308C>A | D3681E | 33 | Yes |
| ORF1ab | 12053C>T | L3930F | 33 | Yes |
| ORF1ab | 14408C>T | P4715L | 33 |  |
| ORF1ab | 17339C>T | A5692V | 32 | Yes |
| S | 21981_21989del | LGV141_143del | 32 | Yes |
| S | 23012G>A | E484K | 33 | Yes |
| S | 23063A>T | N501Y | 33 | Yes |
| S | 23403A>G | D614G | 33 |  |
| S | 23604C>A | P681H | 33 | Yes |
| S | 24836G>A | E1092K | 33 | Yes |
| S | 24863C>T | H1101Y | 33 | Yes |
| S | 25088G>T | V1176F | 33 | Yes |
| ORF8 | 27897A>C | K2Q | 33 | Yes |
| N | 28881_28882GG>AA | R203K | 33 |  |
| N | 28883G>C | G204R | 33 |  |
| **Synonymous Mutations** |  |  |  |  |
| ORF1ab | 3037C>T | F924F | 33 |  |
| ORF1ab | 7564C>T | S2433S | 33 |  |
| ORF1ab | 12049C>T | N3928N | 33 |  |
| S | 23341T>C | G593G | 33 |  |
| S | 24187T>A | S875S | 33 |  |

Among the three known lineages of concern (*i.e.* B.1.1.7, P.1, B.1.351), the B.1.1.7 viruses have been the most frequently detected so far in the Philippines, majority of which were collected from the main ports of entry among ROFs and in a cluster of cases in the Cordillera Administrative Region (CAR), north of the capital Manila. A handful of the B.1.351 viruses have also been detected, but none of the P.1 lineage as of this writing. Infections from SARS-CoV-2 viruses belonging to the B.1.1.28 lineage appear to be more widespread, with cases detected in six (6) different regions. However, the majority of the B.1.1.28 cases in Central Visayas exhibited the signature mutations described for the emergent variant, designated here as PHL-B.1.1.28 (**Figure 3A**).

**Figure 3.**
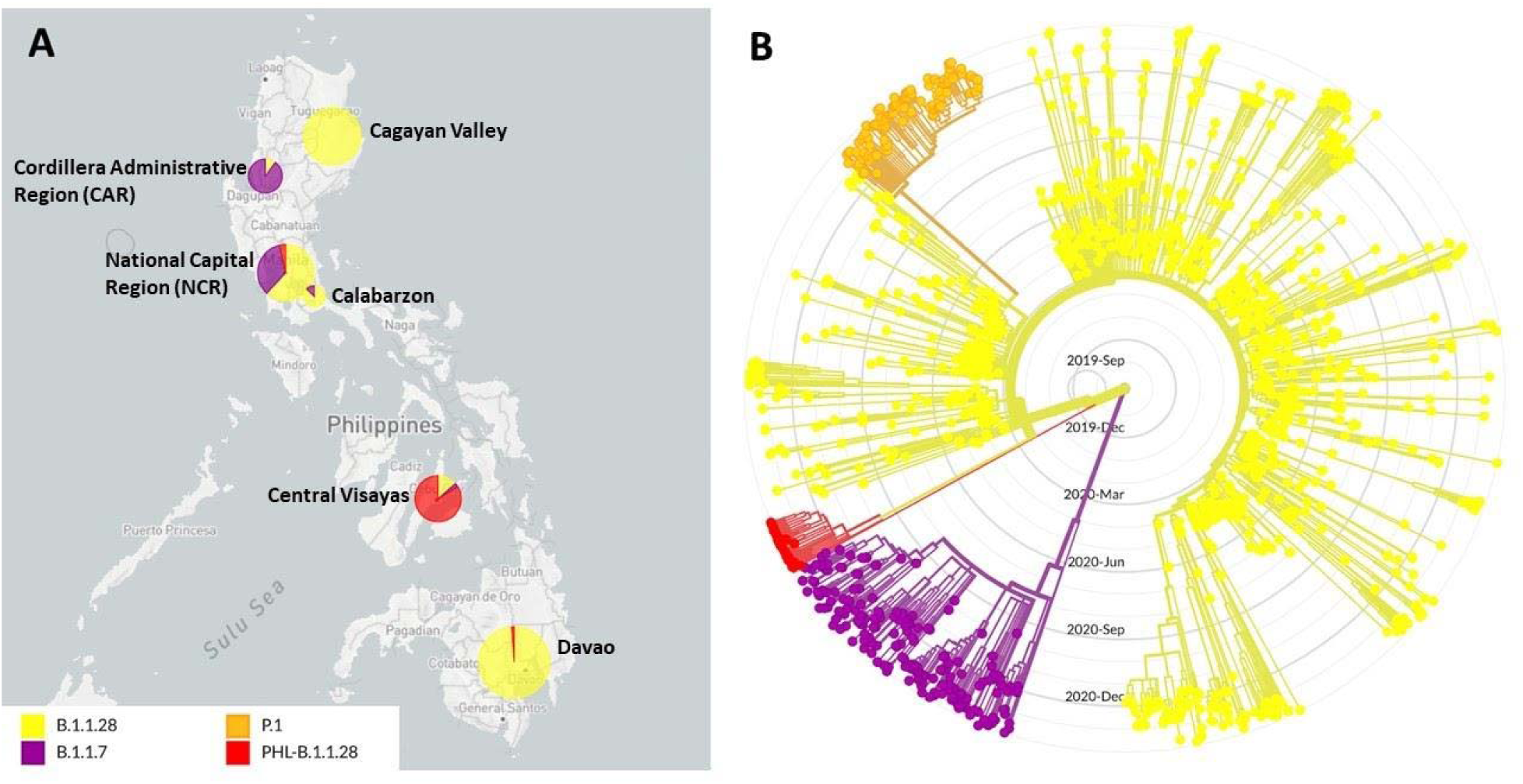
Geographic distribution and phylogenetic clustering of an emergent SARS-CoV-2 lineage in the Philippines designated as PHL-B.1.1.28. **(A)** Geographic distribution of viruses under lineages B.1.1.28 (Yellow), B.1.1.7 (Purple), P.1 (Orange) and the emergent variant PHL-B.1.1.28 (Red) in the Philippines. Most of the samples detected to have the signature mutations of PHL-B.1.1.28 were from the Central Visayas region. Note that viruses from lineage P.1 have not been detected in the country to date. **(B)** Phylogenetic tree of B.1.1.28, B.1.1.7, P.1, and PHL-B.1.1.28 sequences showing that the emergent variant forms a monophyletic clade separate from other lineages included in the analysis. The B.1.1.7 and B.1.1.28 clades include samples collected both globally and in the Philippines.

Phylogenetic tree reconstruction of the depicted local SARS-CoV-2 cases, together with representative sequences of the B.1.1.7, B.1.1.28, and P.1 lineages obtained from GISAID, shows that PHL-B.1.1.28 viruses form a distinct monophyletic clade that is separate from other lineages (**Figure 3B**).

## DISCUSSION

In this study, we detected 33 Philippine SARS-CoV-2 infections, majority of which were collected from laboratory facilities in Central Visayas, National Capital Region, and Davao Region (**Table 1**), caused by viruses initially classified under lineage B.1.1.28, but with the co-occurrence of the E484K, N501Y, and P681H spike protein mutations. These three (3) mutations have all been identified as defining mutations in lineages that have attracted global concern with evidence of biological significance.

The E484K mutation, which is a defining SNP of the B.1.351 and P.1 lineages, has been identified as an immune escape mutation associated with a loss of neutralizing activity by vaccine-elicited antibodies (Collier et al., 2021). The N501Y mutation, which co-occurs with E484K in the B.1.351 and P.1 lineages and is also found in B.1.1.7 viruses, is associated with higher transmissibility and increased binding affinity to the ACE2 receptor (Leung et al., 2021; Luan et al., 2021). The P681H mutation on the other hand that is found in the B.1.1.7 lineage, is immediately adjacent to the furin cleavage site at the S1/S2 boundary of the spike protein. The interaction of this site with furin and the ACE2 receptor is implicated in viral transmission and systemic infection (Hasan et al., 2020; Wang et al, 2020). However, none of the other defining SNPs for these lineages are found in the Philippine samples, suggesting that these three (3) mutations may have arisen independently. All the samples also harbor the globally prevalent D614G mutation, which has been previously associated with increased transmissibility and higher viral loads (Hou et al., 2020; Lorenzo-Redondo et al., 2020).

The mutation profile (**Figure 2**) for this group of viruses further indicates that they have a characteristic combination of mutations not observed in other known SARS-CoV-2 lineages of concern. Apart from E484K, N501Y, D614G, and P681H, we found three (3) other amino acid replacements located towards the C-terminal end of the spike protein in all of the 33 samples: E1092K, H1101Y, and V1176F (**Table 2**). Although much is unknown about the actual effects of these mutations in terms of viral transmissibility, pathogenicity, and its immunogenicity, they all represent radical amino acid replacements with possible implications in protein structure and activity — E1092K being a change from a negatively charged residue to a positively charged one; the H1101Y is a change from a positively charged residue to a hydrophobic one, and the V1176F from a smaller aliphatic residue to a relatively larger aromatic one. Interestingly, a single amino acid replacement (K2Q) at the N-terminal end of the ORF8 protein product was also observed in all samples, which presents a possible significant change from a positively charged residue to a polar uncharged one.

Moreover, we found a deletion of three (3) amino acids at the spike protein residues 141 to 143 (LGV141_143del) in 32 of the 33 samples included in this report. This deletion is adjacent to the tyrosine residue deletion at the spike protein position 144 (Y144del), which is one of the hallmark mutations of viruses in the B.1.1.7 lineage. Deletions in this region of the spike protein, including Δ144/145 and Δ141-144, have been previously shown to alter the protein’s antigenicity, with one particular monoclonal antibody (4A8 MAb) against SARS-CoV-2 unable to detect mutants harboring the said deletions in a Vero E6 (human kidney epithelial) cell line experiment (McCarthy et al., 2021).

Phylogenetic reconstruction of the 33 Philippine samples with representative samples from the B.1.1.7 and B.1.1.28 lineages, including the P.1 lineage which is derived from B.1.1.28, indicate that they represent an emergent lineage within the global phylogenetic tree (**Figure 3B**). The phylogeny reveals that these samples, referred to here as PHL-B.1.1.28, cluster into a well-defined monophyletic clade (red) that diverges from the B.1.1.28 lineage (yellow). Notably, the tree topology shows that this clade does not derive from the P.1 lineage, indicating that the E484K and N501Y mutations in samples may represent recurrent mutations at these sites, rather than inheritance from the latter. The tree also indicates a recent common ancestor for PHL-B.1.1.28, consistent with the emergence of a novel SARS-CoV-2 variant.

The geographic distribution of the variant described here further supports its position to be separate from the B.1.1.28 lineage. While other samples assigned to B.1.1.28 in the Philippines are relatively abundant in other regions and rare in Central Visayas, the reverse is true for samples bearing the signature mutations of PHL-B.1.1.28 (**Figure 3A**), suggesting that the emergence of this variant took place within a geographically separate area of the Philippines centered in Cebu Island. This observation is particularly notable because the Central Visayas region has been experiencing a sharp spike in COVID-19 cases since early January 2021 according to statistics compiled by the Republic of the Philippines Department of Health (**Figure 1**). One possible cause of this spike could be the increased mobility of people post-holiday season or the emergence of a novel variant with higher transmissibility during this period, consistent with the collection dates for the described samples in late January to early February. Although the earliest collection (January 16, 2021) recorded for this group of viruses was from a sample submitted by a testing laboratory situated at the NCR, we note that hardly any samples were collected from Central Visayas for sequencing prior to January 31, highlighting the fact that much still remains hidden about the evolutionary history of this emergent variant.

## CONCLUSIONS

Presented in this report are several lines of evidence suggesting the emergence of a novel variant of SARS-CoV-2 in the Philippines meriting further investigation, represented by 33 samples collected in areas mainly from Central Visayas. First, all of these samples, collectively referred to here as PHL-B.1.1.28, have three (3) spike protein mutations that are considered as defining SNPs for other lineages of concern, and are associated with changes to viral behavior such as increased transmissibility or antigenic escape. Second, they exhibit a characteristic mutation profile not found in other lineages, comprising 13 signature non-synonymous mutations including others in the spike protein that are also implicated in altered protein structure or activity. Third, the samples cluster into a distinct monophyletic clade within the global tree that diverges from B.1.1.28, similar to the P.1 lineage. And fourth, these samples were collected mainly at a time and location that was experiencing a sharp spike in COVID-19 cases, raising the possibility of a more transmissible SARS-CoV-2 variant as a potential cause.

## METHODOLOGY

Nasopharyngeal swabs from SARS-CoV-2 infected individuals were collected and subjected to automated RNA extraction using the PANAMAX™48 Automated Nucleic Acid Extraction Unit with GenAmplify™ BeaDxMax™ Viral DNA/RNA Extraction Kit. Enrichment and sequencing of the extracted SARS-CoV-2 genetic material were subsequently performed using the Illumina COVIDSeq workflow. The resulting sequence reads were mapped to the reference SARS-CoV-2 genome sequence (NCBI Accession No. NC_045512.2) using minimap2 v2.17 (Li, 2018) with preset parameters for accurate genomic short-read mapping (-x sr). The output mapping file was further processed using Samtools v1.10 (Li et al., 2009) to fill the mate coordinates and insert sizes field (fixmate), sort according to reference coordinates (sort), and remove read duplicates (markdup). Mapping and coverage statistics were obtained using Samtools (flagstat and coverage) as well.

The consensus sequence generation and intrahost variant calling were primarily done using the tool iVar v1.3 (Grubaugh et al., 2019) with default parameters. Briefly, primer clipping and quality trimming was implemented using the ivar trim function. Intrahost variants were then detected using a combination of Samtools mpileup and ivar variants, with the parameters described in the iVar manual page (https://andersen-lab.github.io/ivar/html/manualpage.html). Following the same manual, the detected intrahost variants were used to identify and exclude sequence reads associated with mismatched primer indices using the getmasked and removereads functions of iVar. The consensus sequence assemblies were then generated using Samtools mpileup and ivar consensus commands with default parameters (minimum quality = 20, minimum coverage = 10x).

From the consensus sequence assemblies, SARS-CoV-2 lineages were assigned using the tool Pangolin v2.3.0 (https://github.com/cov-lineages/pangolin; Rambaut et al., 2020b). Variants relative to the generated consensus sequences were also identified using MUMmer v4.0 (Kurtz et al., 2004) as implemented in the annotation tool RATT (Otto et al., 2011). Summary mutation tables and plots were generated using in-house scripts written in Python.

Phylogenetic trees were produced by initially generating a multiple sequence alignment of the local SARS-CoV-2 consensus sequences using the tool MAFFT v7.407 (Katoh et al., 2002), supplemented with publicly available sequences from the EpiCoV database of the Global Initiative for Sharing All Influenza Data (GISAID) platform (Shu and McCauley, 2017). The resulting alignment was automatically trimmed using trimAl (Capella-Gutierrez et al., 2009). Phylogenetic trees were then reconstructed using a local instance of the Nextstrain analysis platform (Hadfield et al., 2018).

## Data Availability

Genome sequences of the 33 Philippine SARS-CoV-2 samples reported here are deposited at the EpiCoVTM database of the Global Initiative for Sharing All Influenza Data (GISAID) with accession codes EPI_ISL_1122426 to EPI_ISL_ 1122458.

## ACKNOWLEDGEMENT

This study was funded by the Republic of the Philippines Department of Health, the Philippine Council for Health Research and Development of the Department of Science and Technology, and the University of the Philippines System. The authors would also like to thank the contributing institutions to the Philippine Genomic Biosurveillance Network for their logistical support.

## ETHICS APPROVAL

The methods used in this study has been approved by the Single Joint Research Ethics Board of the Department of Health, Republic of the Philippines, with approval code SJREB-2021-11, as part of a larger research program entitled “A retrospective study on the national genomic surveillance of COVID-19 transmission in the Philippines by SARS-CoV-2 genome sequencing and bioinformatics analysis”.

## DATA AVAILABILITY

Genome sequences of the 33 Philippine SARS-CoV-2 samples reported here are deposited at the EpiCoV™ database of the Global Initiative for Sharing All Influenza Data (GISAID) with accession codes EPI_ISL_1122426 to EPI_ISL_ 1122458. The acknowledgement table for other GISAID sequences used in this study can be found in **Supplementary Table S1**.

